# Emergence in Southern France of a new SARS-CoV-2 variant of probably Cameroonian origin harbouring both substitutions N501Y and E484K in the spike protein

**DOI:** 10.1101/2021.12.24.21268174

**Authors:** Philippe Colson, Jérémy Delerce, Emilie Burel, Jordan Dahan, Agnès Jouffret, Florence Fenollar, Nouara Yahi, Jacques Fantini, Bernard La Scola, Didier Raoult

## Abstract

SARS-CoV-2 variants have become a major virological, epidemiological and clinical concern, particularly with regard to the risk of escape from vaccine-induced immunity. Here we describe the emergence of a new variant. For twelve SARS-CoV-positive patients living in the same geographical area of southeastern France, qPCR testing that screen for variant-associated mutations showed an atypical combination. The index case returned from a travel in Cameroon. The genomes were obtained by next-generation sequencing with Oxford Nanopore Technologies on GridION instruments within ≈8 h. Their analysis revealed 46 mutations and 37 deletions resulting in 30 amino acid substitutions and 12 deletions. Fourteen amino acid substitutions, including N501Y and E484K, and 9 deletions are located in the spike protein. This genotype pattern led to create a new Pangolin lineage named B.1.640.2, which is a phylogenetic sister group to the old B.1.640 lineage renamed B.1.640.1. Both lineages differ by 25 nucleotide substitutions and 33 deletions. The mutation set and phylogenetic position of the genomes obtained here indicate based on our previous definition a new variant we named “IHU”. These data are another example of the unpredictability of the emergence of SARS-CoV-2 variants, and of their introduction in a given geographical area from abroad.

---

SARS-CoV-2 has emerged in China in December 2019 and has been declared a pandemic 21 months ago [1]. We have shown since the summer of 2020 that several SARS-CoV-2 variants have emerged in our geographical area and caused distinct epidemics, either successive or superimposed [2,3]. In addition, we described that the origin of these variants was often their introduction from abroad but could also be mink. This was observed by genotyping, as of 09/12/2021, SARS-CoV-2 from almost 40,000 patients using next-generation sequencing (NGS) of complete genomes for more than 22,000 patients and implementing multiple qPCR specific of each variant for a more exhaustive assessment of their spread. Since then and with the emergence of the Alpha variant at the end of 2020, SARS-CoV-2 variants have become a major virological, epidemiological, and clinical concern, particularly with regard to the risk of escape from vaccine-induced immunity [4-7]. Here we describe the emergence in south-eastern France of a new variant of probably Cameroonian origin.

The index case was an adult first diagnosed as infected with SARS-CoV-2 by real-time reverse transcription PCR (qPCR) performed in a private medical biology laboratory on a nasopharyngeal sample collected mid-November 2021 (Table 1). He was vaccinated against SARS-CoV-2 and returned from a travel to Cameroon three days before. He developed mild respiratory symptoms the day before diagnosis. He lives in a small town of southeastern France. Subsequent detection by qPCR of three mutations in the spike gene to screen for variants, as systematically performed in France in case of SARS-CoV-2 positivity, revealed an atypical combination with L452R-negativity, E484K-positivity, and E484Q-negativity (Pentaplex assay, ID Solution, France) that did not correspond to the pattern of the Delta variant involved in almost all SARS-CoV-2 infections at that time (Table 1). Respiratory samples collected from seven other SARS-CoV-2-positive patients living in the same geographical area exhibited the same combination of mutations screened by qPCR. They were two adults and five children (<15 years of age) (Table 1). The respiratory samples from these eight patients were sent to university hospital institute Méditerranée Infection for SARS-CoV-2 genome sequencing as recommended by French public health authorities. A rapid NGS procedure was launched overnight. It allowed obtaining SARS-CoV-2 genotype identification in ≈8 hours. Briefly, viral RNA was extracted from 200 µL of nasopharyngeal swab fluid using the KingFisher Flex system (Thermo Fisher Scientific, Waltham, MA, USA) following the manufacturer’s instructions. Extracted RNA was reverse-transcribed using SuperScript IV (Thermo Fisher Scientific) and cDNA second strand was synthesized with LunaScript RT SuperMix kit (New England Biolabs) then amplified using a multiplex PCR protocol according to the ARTIC procedure (https://artic.network/) with ARTIC nCoV-2019 V3 panel of primers (IDT, Coralville, IA, USA). Finally, NGS was performed with the ligation sequencing kit and a GridION instrument of Oxford Nanopore Technologies (Oxford, UK) following manufacturer’s instructions. Subsequently, fastq files were processed using the ARTIC field bioinformatics pipeline (https://github.com/artic-network/fieldbioinformatics). NGS reads were basecalled using Guppy (4.0.14) and aligned to the Wuhan-Hu-1 reference genome GenBank accession no. MN908947.3 using minimap2 (v2.17-r941) (https://github.com/lh3/minimap2) [8]. The ARTIC tool align_trim was used to softmask primers from read alignment and to cap sequencing depth at a maximum of 400. The identification of consensus-level variant candidates was performed using the Medaka (0.11.5) workflow developed by ARTIC (https://github.com/artic-network/artic-ncov2019). This strategy allowed assemblying the complete genome from NGS reads obtained within 30 min of run for cycle threshold values (Ct) of qPCR comprised between 15 and 27. SARS-CoV-2 genomes were classified into Nextclade and Pangolin lineages using web applications (https://clades.nextstrain.org/;https://cov-lineages.org/pangolin.html) [10,11,13]. They were deposited in the GISAID sequence database (https://www.gisaid.org/) [14] (Table 1). Phylogenies were reconstructed with the nextstrain/ncov tool (https://github.com/nextstrain/ncov) then visualized with Auspice (https://docs.nextstrain.org/projects/auspice/en/stable/).

**Table 1.**
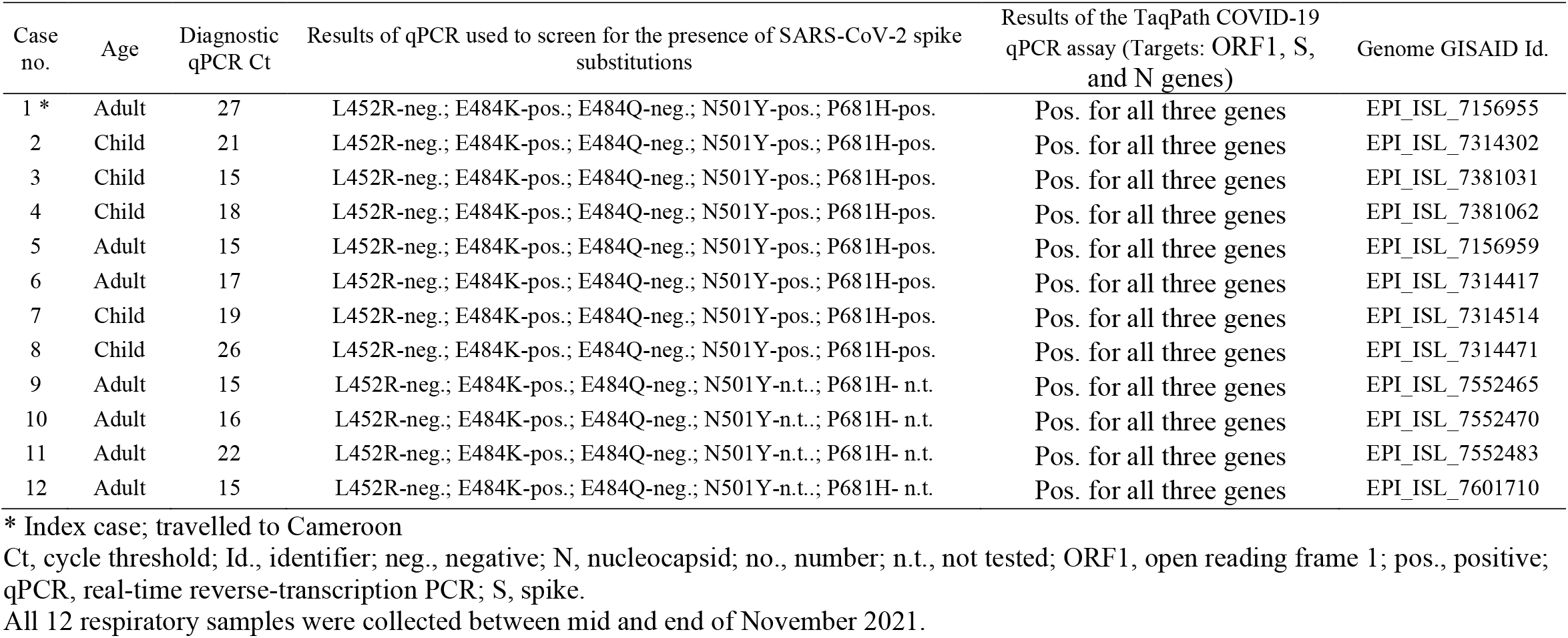
Main epidemiological and virological features of cases identified with infection with the SARS-CoV-2 IHU variant.

The analysis of viral genomes revealed the presence of 46 nucleotide substitutions and 37 deletions, resulting in 30 amino acid substitutions and 12 deletions (Figure 1a; Supplementary Tables S1 and S2). Fourteen amino acid substitutions and 9 amino acid deletions are located in the spike protein. Substitutions N501Y and E484K are combined as in the Beta, Gamma, Theta and Omicron variants [5,15]. Substitution F490S is present as in the Lambda variant, and substitution P681H is present as in the Lambda and Omicron variants. In other structural proteins than the spike, amino acid changes include two substitutions in the nucleocapsid protein and one in the membrane protein. In non-structural proteins, amino acid changes include one substitution in proteins Nsp2, Nsp3, Nsp4, Nsp6, Nsp12 (RNA-dependent RNA polymerase), and Nsp13 (helicase); two substitutions in Nsp14 (3’-5’exonuclease); four substitutions in Nsp8 (which is part of the replication complex with Nsp7 and Nsp12); and three deletions in Nsp6. Finally, in regulatory proteins, amino acid changes include four substitutions in ORF3a, one in ORF9b and one in ORF8. In addition, codon 27 of ORF8 gene is changed into a stop codon, as in the Alpha variant [16]; some members of the Marseille-4 variant (B.1.160) that predominated in our geographical area between August 2020 and February 2021 also exhibit a stop codon in ORF8 gene but at another position [3].

**Figure 1.**
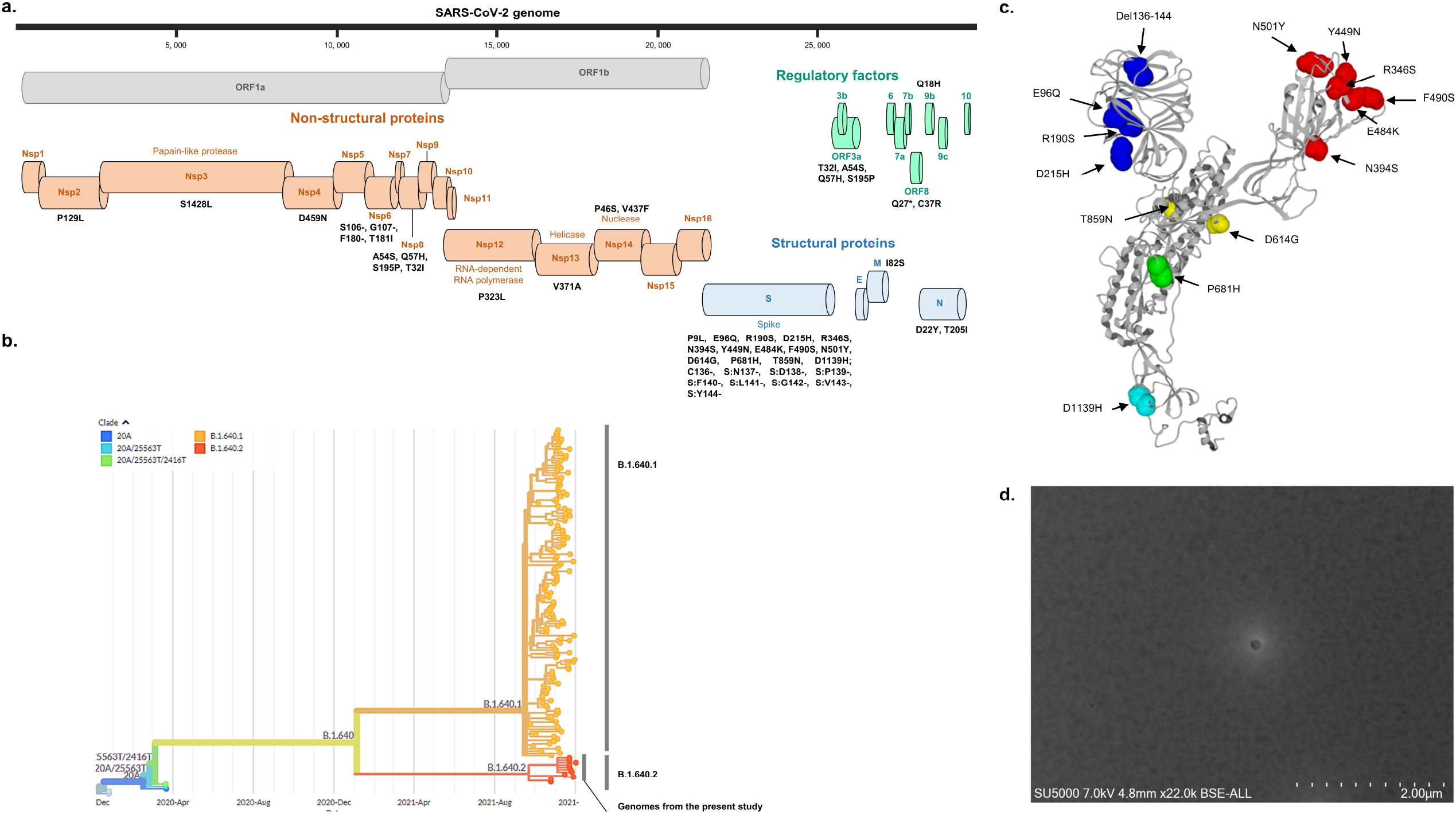
Virological features and scanning electron microscopy image of the SARS-CoV-2 IHU variant. a: Map of the IHU variant genome showing amino acid substitutions and deletions. b: Phylogeny reconstruction performed using the nextstrain/ncov tool (https://github.com/nextstrain/ncov) then visualized with Auspice (https://docs.nextstrain.org/projects/auspice/en/stable/). The genome of the original Wuhan-Hu-1 SARS-CoV-2 isolate (GenBank accession no. NC_045512.2) was added as outgroup, in addition to SARS-CoV-2 genomes of Pangolin lineages B.1.640.1 and B.1.640.2. X-axis shows time. c: Representations of the spike of the IHU variant showing the location of all its amino acid substitutions. N-terminal domain (NTD) mutations are in blue; receptor binding domain (RBD) mutations are in red; mutations involved in ACE-2 unmasking are in yellow; mutations at S1-S2 cleavage site are in green; mutations at fusion region are in cyan. d: Scanning electron microscopy image obtained using a SUV 5000 microscope from a respiratory sample positive for the SARS-CoV-2 IHU variant (Hitachi High-Technologies Corporation, Tokyo, Japan).

Nextclade identified a 20A lineage. Pangolin identified a B.1.640 lineage in primary analysis but a B.1 lineage with the -usher (Ultrafast Sample placement on Existing tRee; https://genome.ucsc.edu/cgi-bin/hgPhyloPlace) option, which showed the phylogenetic placement of the genomes we obtained as an outgroup of the B.1.640 lineage and their clustering with a genome obtained late October in France (Ile-de-France) (EPI_ISL_5926666). The B.1.640 lineage corresponds to a variant first identified in France in April 2021, in Indonesia in August 2021, and in Republic of the Congo (Brazzaville) in September 2021, and it was involved in a cluster of cases in Brittany, France around mid-October 2021 [17]. As of 09/12/2021, 157 genomes were available from the GISAID database including 92 from France and 36 from the Republic of the Congo. The sets of spike mutations of the B.1.640 lineage and of genomes obtained here are similar, with 11 common nucleotide substitutions and 1 common deletion of 9 codons (Supplementary Figure S1, Tables S1-2). However, spike genes of both lineages differ by 7 mutations. In addition, 25 nucleotide substitutions and 33 nucleotide deletions located elsewhere in the genomes differ between the two genotypes. The pattern of mutations of present genomes hence indicates a new variant, which we named “IHU” (in reference to our institute), based on our previous definition [3]. Phylogeny performed with nextstrain/ncov tool (https://github.com/nextstrain/ncov) also showed that B.1.640 and IHU variants were most closely related between each other but comprised two divergent branches (Figure 1b). Their last common ancestor is estimated to date from January 2021 but no genome is currently available from GISAID that corresponds to it. Accordingly, a new Pangolin clade corresponding to the IHU variant was created on 07/12/2021 that was named B.1.640.2, the old B.1.640 clade being renamed B.1.640.1 (https://github.com/cov-lineages/pango-designation/issues/362). It encompasses present genomes and three other genomes comprising a sister group including the one recovered late October 2021 in France (Ile-de-France) (EPI_ISL_5926666) and two additional genomes obtained from samples collected late November in England (EPI_ISL_7181977) and Wales (EPI_ISL_7402094). As the index case was probably infected with the IHU variant during his stay in Cameroon, we sought for this variant in GISAID among genomes from this country but as of 09/12/2021 none of the 556 available genomes belong to the B.1.640.1 or B.1.640.2 lineages.

We analyzed a complete structure of the spike protein of the IHU variant generated by incorporating its specific mutational profile to the original 20B SARS-CoV-2 (Wuhan-Hu-1 isolate with D614G substitution) [18] and fixing all gaps in the pdb file by incorporating the missing amino acids with the Robetta protein structure prediction tool [https://robetta.bakerlab.org/], followed by energy minimization with the Polak-Ribière algorithm as previously reported (Figure 1c) [19]. In the N-terminal domain (NTD), the 134-145 amino acid deletion is predicted to significantly affect the neutralizing epitope. Other changes involve amino acids at positions 96 and 190: in Wuhan-Hu-1 isolate, E96 and R190 induce a turn in NTD secondary structure through electrostatic interactions between each other. This interaction is conserved between substituted amino acids 96Q and 190S, which suggests the co-evolution of these changes. In the receptor binding domain (RBD), aside the well-known substitutions N501Y and E484K, several changes were predicted to significantly affect the neutralizing epitopes. Particularly, P681H is located in the cleavage site of S1-S2 subunits of the spike and is observed in other variants including the recently emerging Omicron [15]. Besides, D1139H substitution implies an amino acid involved in the fusion between the virus and the infected cell.Also, D614G is combined with T859N in the IHU variant. Interestingly, in the Wuhan-Hu-1 isolate, amino acids D614 and T859 from two subunits of the trimeric spike are face to face and lock the trimer in a closed conformation. Substitution D614G allows unlocking the trimer conformation, but this is predicted to be still easier in case of additional presence of substitution T859N.

Respiratory samples collected until end of November 2021 from four other SARS-CoV-2 positive patients living in the same city or borough than the index case could be identified as containing the IHU variant by NGS within 24 hours after their reception (Table 1). All 12 IHU variant-positive samples showed the same combination of spike mutations as screened by real-time qPCR techniques: negativity for 452R and 484Q; positivity for 484K, 501Y [20], and 681H [3]. We also used the TaqPath COVID-19 kit (Thermo Fisher Scientific, Waltham, USA) that provided positive signals for all three genes targeted (ORF1, S, and N). Thus, the IHU variant can be distinguished by screening with qPCR assays from the Delta (L452R-positive) and Omicron (L452R-negative and negative for S gene detection by the TaqPath COVID-19 assay) variants that currently co-circulate in our geographical area. Finally, scanning electron microscopy using a SUV 5000 microscope (Hitachi High-Technologies Corporation, Tokyo, Japan) [21] allowed a quick visualization of the virus from a respiratory sample (Figure 1d).

Overall, these observations show once again the unpredictability of the emergence of new SARS-CoV-2 variants and their introduction from abroad, and they exemplify the difficulty to control such introduction and subsequent spread. They also warrant the implementation of genomic surveillance of SARS-CoV-2 that we started from the very beginning of the pandemic in our geographical area as soon as we diagnosed the first SARS-CoV-2 infection [21] and that we expanded during summer 2020 [2,3]. This surveillance has been implemented at the country scale in 2021 through the French Emergen consortium (https://www.santepubliquefrance.fr/dossiers/coronavirus-covid-19/consortium-emergen). It is too early to speculate on virological, epidemiological or clinical features of this IHU variant based on these 12 cases. For this purpose, respiratory samples from infected patients were inoculated on Vero E6 cells as previously described [22] to be able assessing the sensibility to neutralization by anti-spike antibodies elicited by vaccine immunization, or by prior infection [23].

## Data Availability

All data produced are available online at https://www.gisaid.org.

## Acknowledgments

We are thankful to the Emergen French consortium (https://www.santepubliquefrance.fr/dossiers/coronavirus-covid-19/consortium-emergen). We are also grateful to Laurence Thomas, Claudia Andrieu, Ludivine Brechard, Mamadou Beye, Marielle Bedotto, Elsa Prudent, Sofiane Bakour, Jacques Bou Khalil, and Clio Grimaldier for their technical help.

## Author contributions

Conceived and designed the experiments: PC, DR, JF, BLS. Contributed materials/analysis tools: PC, JDe, EB, JDa, AJ, FF, NY, JF. Analyzed the data: PC, DR, BLS, JD, EB, JF, NY. Wrote the paper: PC, JF, DR. All authors approved the last version of the manuscript.

## Ethics

This study has been approved by the ethics committee of University Hospital Institute (IHU) Méditerranée Infection (N°2021-029). Access to the patients’ biological and registry data issued from the hospital information system was approved by the data protection committee of Assistance Publique-Hôpitaux de Marseille (APHM) and was recorded in the European General Data Protection Regulation registry under number RGPD/APHM 2019-73.

## SUPPLEMENTARY MATERIAL

### SUPPLEMENTARY FIGURE LEGENDS

**Supplementary Figure S1.**
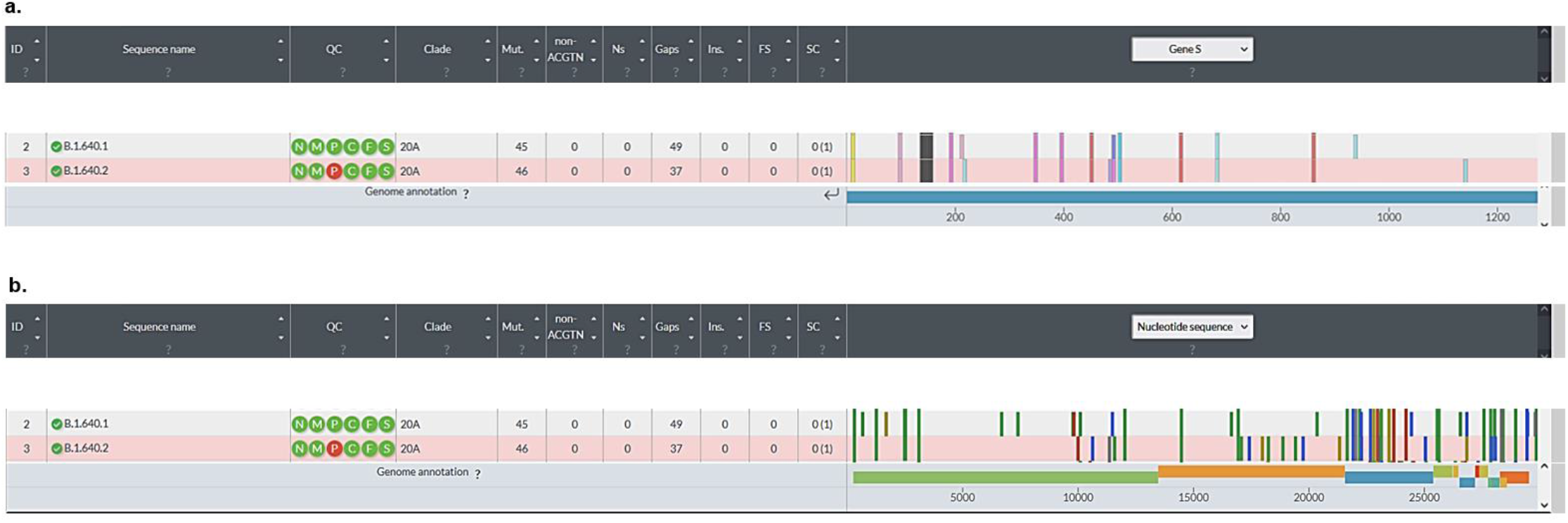
Microarray showing the distribution along the SARS-CoV-2 genome and in viral genes of nucleotide changes observed in comparison with the genome of the Wuhan-Hu-1 isolate for the Pangolin B.1.640.1 and B.1.640.2 (IHU variant) lineages.

Genomes were analyzed using the Nextstrain web-tool (https://clades.nextstrain.org/) [1,2]. Representation is adapted from Nextclade sequence analysis web application output (https://clades.nextstrain.org/).

### SUPPLEMENTARY TABLES

**Supplementary Table S1.** Comparison of nucleotide mutational patterns of the B.1.640.1 and B.1.640.2 lineages.

| B.1.640.1 | B.1.640.2 (IHU variant) |
| --- | --- |
| C241T | C241T |
| C601T | - |
| C1191T | C1191T |
| A1620G | - |
| C2416T | C2416T |
| C2455T | C2455T |
| C3037T | C3037T |
| G6622T | - |
| G7328T | - |
| C9711T | - |
| G9756A | - |
| - | G9929A |
| C10029T | - |
| - | T10561C |
| - | Deletion 11288-11296 |
| T11418C | - |
| - | C11514T |
| C11956T | C11956T |
| C14408T | C14408T |
| C16575T | - |
| C16869T | C16869T |
| - | C17004T |
| - | T17348C |
| - | A17916G |
| - | C18175T |
| - | C18804T |
| - | G19348T |
| - | T19680C |
| C20283T | - |
| - | A21258G |
| C21588T | C21588T |
| G21848C | G21848C |
| Deletion 21968-21994 | Deletion 21968-21994 |
| G22132T | G22132T |
| T22191C | - |
| - | G22205C |
| A22600C | A22600C |
| A22743G | A22743G |
| T22907A | T22907A |
| - | G23012A |
| T23030C | - |
| T23031G | T23031C |
| A23063T | A23063T |
| A23403G | A23403G |
| C23604A | C23604A |
| C24138A | C24138A |
| G24368C | - |
| - | G24977C |
| C25487T | C25487T |
| G25563T | G25563T |
| A26492T | - |
| T26767C | T26767G |
| C27513T | C27513T |
| C27807T | C27807T |
| - | T27833C |
| C27972T | C27972T |
| - | T28002C |
| Deletion 28271 | Deletion 28271 |
| T28297C | C28312T |
| G28337T | G28337T |
| C28887T | C28887T |
| T29377C | T29377C |
| G29405C | - |
| Deletion 29738-29758 | - |
| G29779T | G29779T |

Genomes were analyzed using the Nextstrain web-tool (https://clades.nextstrain.org/) [1,2]. Spike region is indicated by a grey background.

**Supplementary Table S2.** Comparison of amino acid mutational patterns of the B.1.640.1 and B.1.640.2 lineages.

| B.1.640.1 | B.1.640.2 (IHU variant) |
| --- | --- |
| M:I82T | M:I82S |
| N:D22Y | N:D22Y |
| N:T205I | N:T205I |
| N:E378Q | - |
| ORF1a:P309L | ORF1a:P309L |
| ORF1a:E452G | - |
| ORF1a:L2119F | - |
| ORF1a:A2355S | - |
| ORF1a:S3149F | - |
| ORF1a:R3164H | - |
| - | ORF1a:D3222N |
| ORF1a:T3255I | - |
| - | Deletions ORF1a:S3675-, ORF1a:G3676-,<br>ORF1a:F3677- |
| ORF1a:V3718A | - |
| - | ORF1a:T3750I |
| ORF1b:P314L | ORF1b:P314L |
| - | ORF1b:V1294A |
| - | ORF1b:P1570S |
| - | ORF1b:V1961F |
| ORF3a:T32I | ORF3a:T32I |
| ORF3a:Q57H | ORF3a:Q57H |
| ORF8:Q27* | ORF8:Q27* |
| ORF9b:I5T | - |
| ORF9b:Q18H | - |
| - | ORF8:C37R |
| - | ORF9b:P10L |
| - | ORF9b:Q18H |
| S:P9L | S:P9L |
| S:E96Q | S:E96Q |
| Deletions S:C136-, S:N137-, S:D138-,<br>S:P139-, S:F140-, S:L141-, S:G142-,<br>S:V143-, S:Y144- | Deletions S:C136-, S:N137-, S:D138-,<br>S:P139-, S:F140-, S:L141-, S:G142-,<br>S:V143-, S:Y144- |
| S:R190S | S:R190S |
| S:I210T | - |
| - | S:D215H |
| S:R346S | S:R346S |
| S:N394S | S:N394S |
| S:Y449N | S:Y449N |
| - | S:E484K |
| S:F490R | S:F490S |
| S:N501Y | S:N501Y |
| S:D614G | S:D614G |
| S:P681H | S:P681H |
| S:T859N | S:T859N |
| S:D936H | - |
| - | S:D1139H |

Genomes were analyzed using the Nextstrain web-tool (https://clades.nextstrain.org/) [1,2]. Spike region is indicated by a grey background.

